# Association between SARS-CoV-2 Infection and Select Symptoms and Conditions 31 to 150 Days After Testing among Children and Adults

**DOI:** 10.1101/2022.12.18.22283646

**Authors:** Yongkang Zhang, Alfonso Romieu-Hernandez, Tegan K. Boehmer, Eduardo Azziz-Baumgartner, Thomas W. Carton, Adi V. Gundlapalli, Julia Fearrington, Kshema Nagavedu, Katherine Dea, Erick Moyneur, Lindsey G. Cowell, Rainu Kaushal, Kenneth H. Mayer, Jon Puro, Sonja A. Rasmussen, Deepika Thacker, Mark G. Weiner, Sharon Saydeh, Jason P. Block, PCORnet Network Partners

**Author notes:** **Corresponding author** Jason P. Block. Contributed equally (Sharon Saydeh, Jason P. Block). Collaborative authors listed in the Acknowledgement. **IRB Approval:** This cohort study, included within a general SARS-CoV-2 surveillance project across PCORnet institutions, was deemed exempt from review under the public health surveillance provision of the Common Rule by the Harvard Pilgrim Health Care institutional review board. **Disclaimer** The findings and conclusions in this report are those of the author(s) and do not necessarily represent the official position of the Centers for Disease Control and Prevention (CDC), the RECOVER Program, or the National Institutes of Health.

## Abstract

**Background:** An increasing number of studies have described new and persistent symptoms and conditions as potential post-acute sequelae of SARS-CoV-2 infection (PASC). However, it remains unclear whether certain symptoms or conditions occur more frequently among persons with SARS-CoV-2 infection compared with those never infected with SARS-CoV-2. We compared the occurrence of specific COVID-associated symptoms and conditions as potential PASC 31 to 150 days following a SARS-CoV-2 test among adults (≥20 years) and children (<20 years) with positive and negative test results documented in the electronic health records (EHRs) of institutions participating in PCORnet, the National Patient-Centered Clinical Research Network.

**Methods and Findings:** This study included 3,091,580 adults (316,249 SARS-CoV-2 positive; 2,775,331 negative) and 675,643 children (62,131 positive; 613,512 negative) who had a SARS-CoV-2 laboratory test (nucleic acid amplification or rapid antigen) during March 1, 2020–May 31, 2021 documented in their EHR. We identified hospitalization status in the day prior through the 16 days following the SARS-CoV-2 test as a proxy for the severity of COVID-19. We used logistic regression to calculate the odds of receiving a diagnostic code for each symptom outcome and Cox proportional hazard models to calculate the risk of being newly diagnosed with each condition outcome, comparing those with a SARS-CoV-2 positive test to those with a negative test. After adjustment for baseline covariates, hospitalized adults and children with a positive test had increased odds of being diagnosed with ≥1 symptom (adults: adjusted odds ratio[aOR], 1.17[95% CI, 1.11-1.23]; children: aOR, 1.18[95% CI, 1.08-1.28]) and shortness of breath (adults: aOR, 1.50[95% CI, 1.38-1.63]; children: aOR, 1.40[95% CI, 1.15-1.70]) 31-150 days following a SARS-CoV-2 test compared with hospitalized individuals with a negative test. Hospitalized adults with a positive test also had increased odds of being diagnosed with ≥3 symptoms (aOR, 1.16[95% CI, 1.08 – 1.26]) and fatigue (aOR, 1.12[95% CI, 1.05 – 1.18]) compared with those testing negative. The risks of being newly diagnosed with type 1 or type 2 diabetes (aHR, 1.25[95% CI, 1.17-1.33]), hematologic disorders (aHR, 1.19[95% CI, 1.11-1.28]), and respiratory disease (aHR, 1.44[95% CI, 1.30-1.60]) were higher among hospitalized adults with a positive test compared with those with a negative test. Non-hospitalized adults with a positive SARS-CoV-2 test had higher odds of being diagnosed with fatigue (aOR, 1.11[95% CI, 1.05-1.16]) and shortness of breath (aOR, 1.22[95% CI, 1.15-1.29]), and had an increased risk (aHR, 1.12[95% CI, 1.02-1.23]) of being newly diagnosed with hematologic disorders (i.e., venous thromboembolism and pulmonary embolism) 31-150 days following SARS-CoV-2 test compared with those testing negative. The risk of being newly diagnosed with certain conditions, such as mental health conditions and neurological disorders, was lower among patients with a positive viral test relative to those with a negative viral test.

**Conclusions:** Patients with SARS-CoV-2 infection were at higher risk of being diagnosed with certain symptoms and conditions, particularly fatigue, respiratory symptoms, and hematological abnormalities, after acute infection. The risk was highest among adults hospitalized after SARS-CoV-2 infection.

## Introduction

Studies have reported that 10-50% of individuals infected with SARS-CoV-2 develop new and persistent symptoms and conditions after the acute infection [1-4]. These new symptoms and conditions, sometimes referred to as post-acute sequelae of SARS-CoV-2 infection (PASC) or long COVID, affect a wide range of organ systems [5, 6]. Previous studies have identified new onset of fatigue or muscle weakness [1, 3, 7-9], shortness of breath [1, 8-10], cognitive dysfunction [8, 11, 12], pulmonary diseases [3], cardiovascular diseases [11], diabetes [3], mental health conditions [3, 8, 10], and adverse kidney outcomes [3, 13] as among the most common PASC. Studies also have found that the occurrence of PASC is not uniform, with higher incidence among those who had higher severity of the acute SARS-CoV-2 infection (e.g., hospitalized or requiring invasive mechanical ventilation) and older age, among other patient subgroups [14, 15].

Understanding the symptoms and conditions specifically associated with SARS-CoV-2 infection is critical to help guide clinical monitoring and treatment along with public health response and resource allocation to PASC. However, significant gaps still exist about our understanding of new symptoms and conditions following SARS-CoV-2 infection. Although a few population-based studies have used large samples to examine PASC symptoms or conditions, they only focused on specific patient populations, such as US veterans and Medicare patients [1, 3, 14].

Prior studies focusing on a generalizable population of adults have primarily included cohorts of hospitalized patients with COVID-19 without use of a control group, examined PASC symptoms and conditions related to a single organ system, focused on COVID-19 patients from a specific region or from the early waves of the pandemic, or did not adjust for some potential confounders between SARS-CoV-2 infection and PASC symptoms and conditions [7, 16-21]. Research on PASC among children is emerging, with few studies using large cohorts or comparing hospitalized and non-hospitalized patients [22-24]. Symptoms and conditions following SARS-CoV-2 infection among children and non-hospitalized adults also have not been well characterized with large samples. Some studies used patient-reported data collected from surveys or interviews [11, 12, 25, 26]. These data could provide more detailed information about patient experience that is not captured in healthcare data, such as electronic health records. However, these patient-reported data may capture symptoms at only one point in time and may not account for symptoms and conditions before SARS-CoV-2 infection.

In this study, we used electronic health records (EHR) data from health systems participating in PCORnet, the National Patient-Centered Clinical Research Network [27], to examine whether select symptoms and conditions were associated with SARS-CoV-2 infection among adults and children compared with a control population of those who had only negative tests for SARS-CoV-2.

## Methods

### Study Setting

PCORnet is a network of more than 60 participating sites, each representing one or more health systems, that facilitates multi-site research using EHR data [28]. The network utilizes a common data model that fosters interoperability across participating sites. Each site transforms their source clinical data into a standardized common data model, with data elements across most domains available in the EHR, including laboratory tests, diagnoses, procedures, prescriptions, demographics, and vital measures, among other information [27]. Data undergoes quarterly data curation to ensure that data meets designated quality standards of the network. Single statistical programs can then be written to access and analyze the data at each site [29]. These analyses, which are executed in a distributed manner (at the site level), can generate aggregate descriptive data in the form of counts and frequencies or summary results from regression analyses that are returned to a study coordinating center and combined into multi-site aggregated results tables.

This study utilized data from 43 PCORnet sites participating in a national COVID-19 surveillance program funded by the Centers for Disease Control and Prevention (eTable 1). Starting in April 2020, sites have refreshed data at least monthly for a cohort of patients receiving care in their affiliated health systems who had a documented SARS-CoV-2 test (defined by Logical Observation Identifiers Names and Codes [25]), a diagnostic code for a respiratory illness including but not limited to COVID-19 (International Classification of Diseases, Tenth Revision, Clinical Modification, ICD-10-CM [30]), or a code for a COVID-19 vaccine or therapeutic (RxNorm [31], National Drug Code [32], or procedure code [33]). These datasets, kept at participating sites, have comprehensive historical information (e.g., diagnosis, procedures, and prescriptions before SARS-CoV-2 infection) about patients meeting these inclusion criteria, but the data do not include all patients cared for in the health systems of a participating site (e.g., those without a SARS-CoV-2 test or a respiratory illness diagnosis).

### Study Population

This study assessed all patients who had a SARS-CoV-2 laboratory test from March 1, 2020, through May 31, 2021. To be included in the study, patients had to have an encounter within a health system in the 540-to 31-day period prior to (baseline period) and in the 31-to 150-day period after (follow-up period) their index test date. This requirement ensured that patients had some engagement with a health system at baseline, allowing us to identify conditions and symptoms that were new after SARS-CoV-2 infection. This also enabled us to account for other baseline risk factors (e.g., age and baseline comorbidities) for PASC symptoms and conditions. Patients were stratified into a child, adolescent, and young adult cohort (aged 0-19 years, hereafter referred to as “youth cohort” or “children”) and an adult cohort (aged ≥20 years) based on their age at the index test date. Age cohorts were further stratified by their hospitalization status based on the care setting associated with the SARS-CoV-2 laboratory test. Hospitalized cohorts included patients with a hospitalization encounter on the day prior through the 16 days following the index test date. Non-hospitalized cohorts included patients without a hospitalization encounter on the day prior through the 16 days following the index test date; these patients either had an ambulatory or emergency department encounter or an encounter with no care setting specified (presumed to be ambulatory, such as telemedicine or testing only).

### PASC Symptoms and Conditions

From prior studies, including another study using PCORnet data, we identified conditions and symptoms that may be more common among those testing positive for SARS-CoV-2 compared with those testing negative and assessed them in this study [1-4, 7-10]. Conditions included mental health conditions (e.g., anxiety, depression, psychosis), chronic kidney disorders, diabetes mellitus type 1 or 2, hematologic disorders (e.g., venous thromboembolism), major cardiovascular events, neurological disorders (e.g., autonomic disorders, Parkinson’s, seizures, dementia), and respiratory diseases. We identified these conditions using ICD-10-CM diagnosis codes. Because we examined the incidence of these conditions in a narrow time window, we only required one code to be present. We examined these conditions as potential PASC only in the adult cohorts as these conditions are extremely rare among patients aged less than 20 years.

Assessed symptoms included fatigue or muscle weakness, shortness of breath or dyspnea, cough, change in bowel habits, abdominal pain, headache, cognitive disorders, disorders of taste and smell, non-cardiac chest pain, heart rate abnormalities, sleep disorders and myalgias/arthralgias. From this list of symptoms, we created four symptom-related outcomes for both adult and youth cohorts. These included two composite outcomes: 1) at least one symptom, which required only one ICD-10-CM code for any of the symptoms above and 2) three or more symptoms, which required at least 3 ICD-10-CM codes for the same or different symptoms; and two single symptom outcomes: 3) fatigue or muscle weakness and 4) shortness of breath or dyspnea. We examined these single symptom outcomes because they were among the most prevalent symptoms after SARS-CoV-2 infection in our prior study of post-COVID conditions and symptoms [4].

We examined all condition and symptom outcomes in the 31-to-150-day period after the index SARS-CoV-2 test date. The queries distributed to PCORnet participating sites were completed by February 1, 2022 (condition outcomes for adults) and March 29, 2022 (symptom outcomes for both adults and children). As a result, all patients included had the opportunity to have an outcome documented for the entire follow-up period of 31 to 150 days after index test dates between March 1, 2020, through May 31, 2021. For the symptom outcomes, we also examined the period of 90 to 150 days as secondary analyses.

### Exposures and Covariates

The exposure of interest was a positive SARS-CoV-2 test, defined as “positive,” “presumptive positive,” or “detected” (“positive viral test”), versus a negative SARS-CoV-2 test, defined as “negative” or “not detected” (“negative viral test”), on a rapid antigen (1% of patients) or nuclear acid amplification test (NAAT) recorded as polymerase chain reaction (PCR) tests (99% of patients). If patients had any positive SARS-CoV-2 viral test during the study period, they were analyzed as having only a positive test regardless of whether they had prior or subsequent negative tests. Patients categorized as having a negative viral test only had negative viral tests throughout the study period. The index test date from which we examined outcomes was the date of the first positive or negative test.

We controlled for several a priori confounders in our regression analyses. For both children and adults, we controlled for age as a continuous variable, age squared to account for nonlinear effect of age, sex (female, male, and missing sex), race (Asian, Black, White, other race, missing), ethnicity (Hispanic, non-Hispanic, missing), weight class (children: BMI less than the 95^th^ percentile, BMI greater or equal to 95th percentile, missing BMI; adults: BMI < 30 kg/m^2^, ≥ 30 kg/m^2^, missing BMI), and number of visits or encounters with a health system in the 150-to 31-day period prior to the index date. For adults, we additionally controlled for combined comorbidity score [34] assessed based on conditions that occurred in the 540 to 7 days prior to the index date and current smoking status (current smoker; never or missing smoking), assessed based on the record closest to the index date in that same period. For hospitalized adults and children, we additionally controlled for length of stay, dexamethasone use, and mechanical ventilation during the hospitalization to account for variation in disease severity among hospitalized patients. Mechanical ventilation was identified from the index date through 16 days following the index date; this time period was chosen to account for the possibility that it may take more than two weeks for respiratory failure to develop.

### Analyses

All analyses were conducted using distributed regression modeling, in which each site separately executed identical regression models, returning summary output including parameter estimates, standard errors, covariance matrices, convergence status, and number of observations. Based on the convergence of each regression at each site, results were either discarded or included in the meta-analysis. Results from a specific site could be discarded for some outcomes and included for others. Once the convergence was assessed, results from the selected sites were combined using meta-analytic techniques (eTable 2). The random-effects model based on the DerSimonian and Laird method was used to obtain pooled estimates [35].

We used different methodological approaches to examine condition and symptom outcomes. Among adults, we examined each of the seven conditions in separate models. For each model, we excluded all patients who had a diagnostic code for the relevant condition that was the outcome for the model during the 540 to 31 days prior to the index date (e.g., patients with hematologic conditions in the baseline were excluded from the model examining the outcome of hematologic conditions). We used Cox proportional hazard regression models, accounting for time from the beginning of the post-acute period (31 days post) to the earliest documentation of the first diagnostic code for each condition (event) and the end of the outcome period (150 days post-censoring). We controlled for all covariates described above in these models.

For the symptom outcomes, we did not exclude patients who had diagnostic codes for these symptoms during the baseline period. We took this approach because of how common these symptoms are in routine clinical care. Instead, we controlled for the presence of these symptoms in the 150 to 31 days prior to index date. We used logistic regression models to assess the odds of having any of these four symptom outcomes associated with SARS-CoV-2 infection in the entire 31 to 150 days post index period. We controlled for the same covariates as we did in the condition outcome models, with the addition of a covariate indicating the presence of relevant symptoms during the baseline period (e.g., for the “any symptom” outcome, we controlled for the presence of any of the symptoms during the baseline period as one of the covariates; for the fatigue outcome, we controlled for the presence of fatigue during the baseline period as one of the covariates). As a secondary analysis, we examined two symptom outcomes, at least one symptom and 3 or more symptoms, associated with SARS-CoV-2 infection in the 90 to 150 days post index test date using logistic regressions. We included this analysis to determine if the associations between SARS-CoV-2 infection and symptom outcomes were persistent 90 days after testing.

All analyses were done using the most recent version of SAS available at each of the sites executing analyses (Cary, NC). This activity was reviewed by CDC and conducted consistent with applicable federal law and CDC policy.

## Results

### Population Characteristics

During March 1, 2020–May 31, 2021, we identified 3,091,580 adults aged 20 years or older meeting the inclusion criteria. Of these patients, 316,249 had a positive viral test, including 270,441 non-hospitalized and 45,808 hospitalized adults; and 2,775,331 had a negative viral test, including 2,102,408 non-hospitalized and 672,923 hospitalized adults. We also identified 675,643 children 19 years or younger meeting the inclusion criteria. Among these patients, 62,131 had a positive viral test, including 59,374 non-hospitalized and 2,757 hospitalized children; and 613,512 had a negative test, including 520,816 non-hospitalized and 92,696 hospitalized children (Table 1).

**Table 1.**
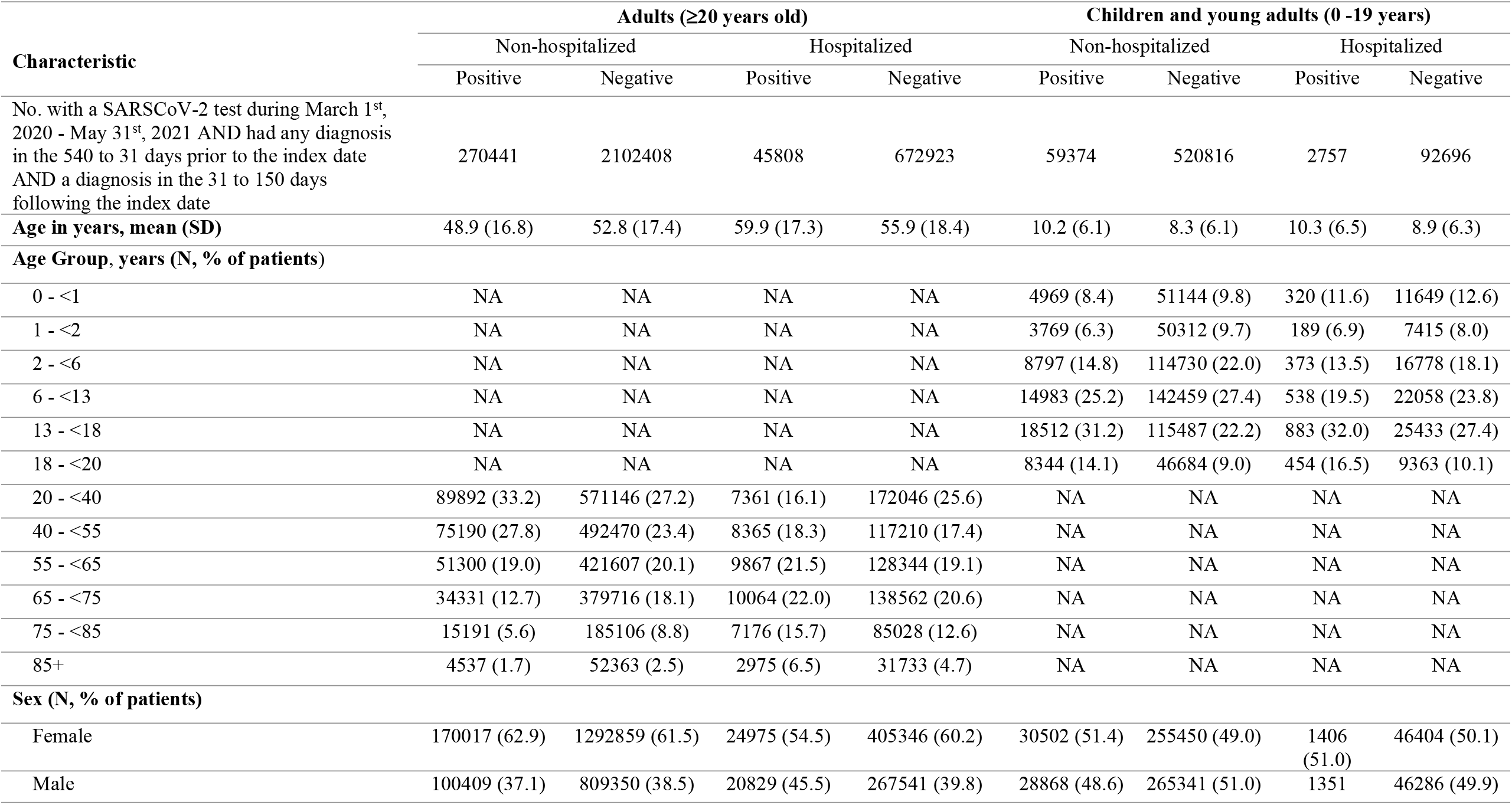
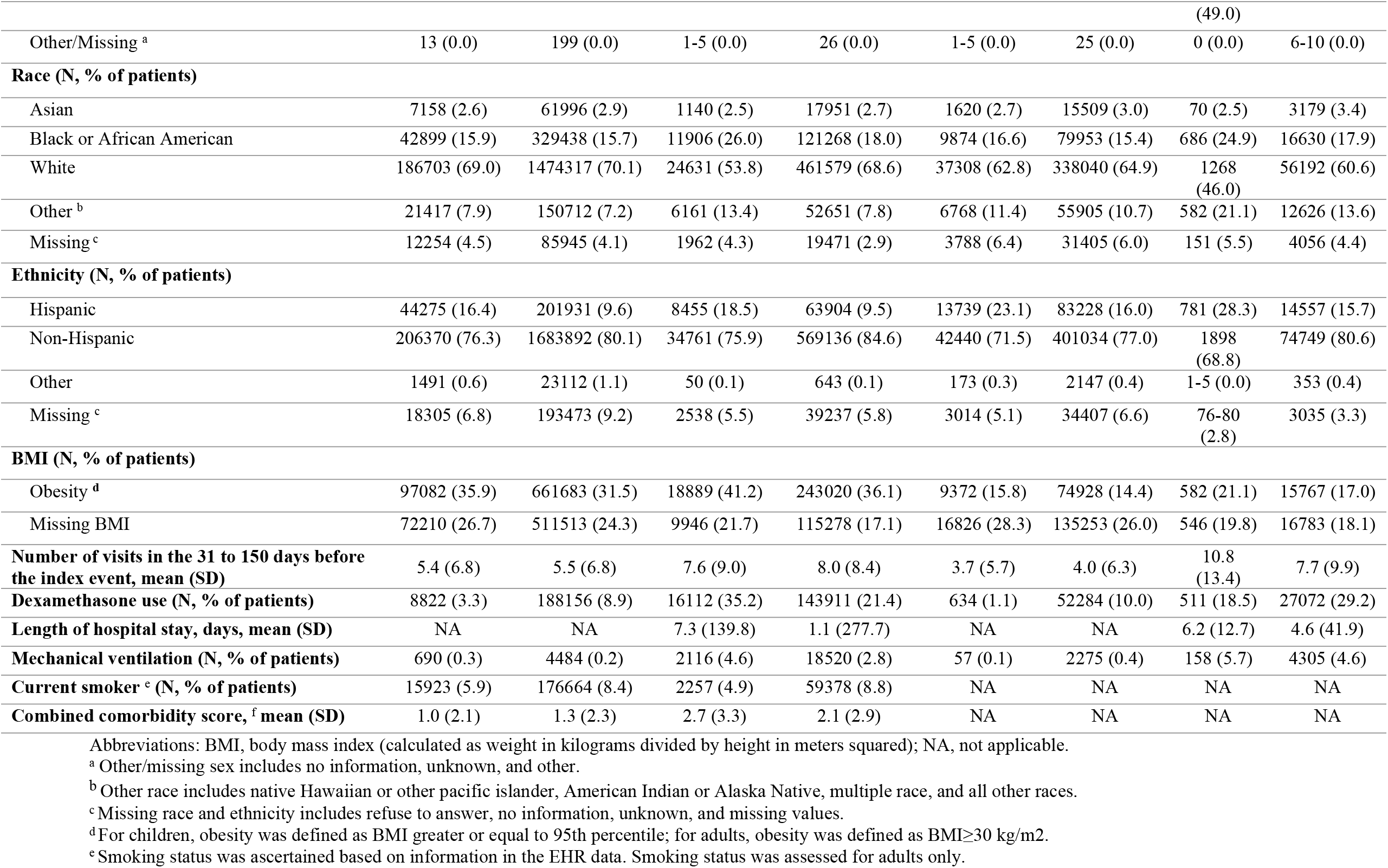
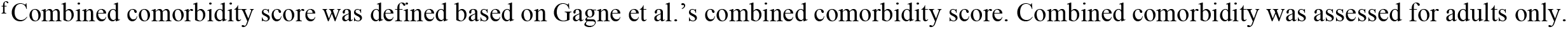
Demographic and Clinical Characteristics of Adults and Children with a Positive or Negative SARS-CoV-2 Test Result.

| Characteristic | Adults (≥20 years old) |  |  |  | Children and young adults (0 -19 years) |  |  |  |
| --- | --- | --- | --- | --- | --- | --- | --- | --- |
|  | Non-hospitalized |  | Hospitalized |  | Non-hospitalized |  | Hospitalized |  |
|  | Positive | Negative | Positive | Negative | Positive | Negative | Positive | Negative |
| No. with a SARSCoV-2 test during March 1<sup>st</sup>, 2020 - May 31<sup>st</sup>, 2021 AND had any diagnosis in the 540 to 31 days prior to the index date AND a diagnosis in the 31 to 150 days following the index date | 270441 | 2102408 | 45808 | 672923 | 59374 | 520816 | 2757 | 92696 |
| **Age in years, mean (SD)** | 48.9 (16.8) | 52.8 (17.4) | 59.9 (17.3) | 55.9 (18.4) | 10.2 (6.1) | 8.3 (6.1) | 10.3 (6.5) | 8.9 (6.3) |
| **Age Group, years (N, % of patients)** |  |  |  |  |  |  |  |  |
| 0 - <1 | NA | NA | NA | NA | 4969 (8.4) | 51144 (9.8) | 320 (11.6) | 11649 (12.6) |
| 1 - <2 | NA | NA | NA | NA | 3769 (6.3) | 50312 (9.7) | 189 (6.9) | 7415 (8.0) |
| 2 - <6 | NA | NA | NA | NA | 8797 (14.8) | 114730 (22.0) | 373 (13.5) | 16778 (18.1) |
| 6 - <13 | NA | NA | NA | NA | 14983 (25.2) | 142459 (27.4) | 538 (19.5) | 22058 (23.8) |
| 13 - <18 | NA | NA | NA | NA | 18512 (31.2) | 115487 (22.2) | 883 (32.0) | 25433 (27.4) |
| 18 - <20 | NA | NA | NA | NA | 8344 (14.1) | 46684 (9.0) | 454 (16.5) | 9363 (10.1) |
| 20 - <40 | 89892 (33.2) | 571146 (27.2) | 7361 (16.1) | 172046 (25.6) | NA | NA | NA | NA |
| 40 - <55 | 75190 (27.8) | 492470 (23.4) | 8365 (18.3) | 117210 (17.4) | NA | NA | NA | NA |
| 55 - <65 | 51300 (19.0) | 421607 (20.1) | 9867 (21.5) | 128344 (19.1) | NA | NA | NA | NA |
| 65 - <75 | 34331 (12.7) | 379716 (18.1) | 10064 (22.0) | 138562 (20.6) | NA | NA | NA | NA |
| 75 - <85 | 15191 (5.6) | 185106 (8.8) | 7176 (15.7) | 85028 (12.6) | NA | NA | NA | NA |
| 85+ | 4537 (1.7) | 52363 (2.5) | 2975 (6.5) | 31733 (4.7) | NA | NA | NA | NA |
| **Sex (N, % of patients)** |  |  |  |  |  |  |  |  |
| Female | 170017 (62.9) | 1292859 (61.5) | 24975 (54.5) | 405346 (60.2) | 30502 (51.4) | 255450 (49.0) | 1406 (51.0) | 46404 (50.1) |
| Male | 100409 (37.1) | 809350 (38.5) | 20829 (45.5) | 267541 (39.8) | 28868 (48.6) | 265341 (51.0) | 1351 | 46286 (49.9) |
|  |  |  |  |  |  |  | (49.0) |  |
| Other/Missing <sup>a</sup> | 13 (0.0) | 199 (0.0) | 1-5 (0.0) | 26 (0.0) | 1-5 (0.0) | 25 (0.0) | 0 (0.0) | 6-10 (0.0) |
| **Race (N, % of patients)** |  |  |  |  |  |  |  |  |
| Asian | 7158 (2.6) | 61996 (2.9) | 1140 (2.5) | 17951 (2.7) | 1620 (2.7) | 15509 (3.0) | 70 (2.5) | 3179 (3.4) |
| Black or African American | 42899 (15.9) | 329438 (15.7) | 11906 (26.0) | 121268 (18.0) | 9874 (16.6) | 79953 (15.4) | 686 (24.9) | 16630 (17.9) |
| White | 186703 (69.0) | 1474317 (70.1) | 24631 (53.8) | 461579 (68.6) | 37308 (62.8) | 338040 (64.9) | 1268 (46.0) | 56192 (60.6) |
| Other <sup>b</sup> | 21417 (7.9) | 150712 (7.2) | 6161 (13.4) | 52651 (7.8) | 6768 (11.4) | 55905 (10.7) | 582 (21.1) | 12626 (13.6) |
| Missing <sup>c</sup> | 12254 (4.5) | 85945 (4.1) | 1962 (4.3) | 19471 (2.9) | 3788 (6.4) | 31405 (6.0) | 151 (5.5) | 4056 (4.4) |
| **Ethnicity (N, % of patients)** |  |  |  |  |  |  |  |  |
| Hispanic | 44275 (16.4) | 201931 (9.6) | 8455 (18.5) | 63904 (9.5) | 13739 (23.1) | 83228 (16.0) | 781 (28.3) | 14557 (15.7) |
| Non-Hispanic | 206370 (76.3) | 1683892 (80.1) | 34761 (75.9) | 569136 (84.6) | 42440 (71.5) | 401034 (77.0) | 1898 (68.8) | 74749 (80.6) |
| Other | 1491 (0.6) | 23112 (1.1) | 50 (0.1) | 643 (0.1) | 173 (0.3) | 2147 (0.4) | 1-5 (0.0) | 353 (0.4) |
| Missing <sup>c</sup> | 18305 (6.8) | 193473 (9.2) | 2538 (5.5) | 39237 (5.8) | 3014 (5.1) | 34407 (6.6) | 76-80 (2.8) | 3035 (3.3) |
| **BMI (N, % of patients)** |  |  |  |  |  |  |  |  |
| Obesity <sup>d</sup> | 97082 (35.9) | 661683 (31.5) | 18889 (41.2) | 243020 (36.1) | 9372 (15.8) | 74928 (14.4) | 582 (21.1) | 15767 (17.0) |
| Missing BMI | 72210 (26.7) | 511513 (24.3) | 9946 (21.7) | 115278 (17.1) | 16826 (28.3) | 135253 (26.0) | 546 (19.8) | 16783 (18.1) |
| **Number of visits in the 31 to 150 days before the index event, mean (SD)** | 5.4 (6.8) | 5.5 (6.8) | 7.6 (9.0) | 8.0 (8.4) | 3.7 (5.7) | 4.0 (6.3) | 10.8 (13.4) | 7.7 (9.9) |
| **Dexamethasone use (N, % of patients)** | 8822 (3.3) | 188156 (8.9) | 16112 (35.2) | 143911 (21.4) | 634 (1.1) | 52284 (10.0) | 511 (18.5) | 27072 (29.2) |
| **Length of hospital stay, days, mean (SD)** | NA | NA | 7.3 (139.8) | 1.1 (277.7) | NA | NA | 6.2 (12.7) | 4.6 (41.9) |
| **Mechanical ventilation (N, % of patients)** | 690 (0.3) | 4484 (0.2) | 2116 (4.6) | 18520 (2.8) | 57 (0.1) | 2275 (0.4) | 158 (5.7) | 4305 (4.6) |
| **Current smoker <sup>e</sup> (N, % of patients)** | 15923 (5.9) | 176664 (8.4) | 2257 (4.9) | 59378 (8.8) | NA | NA | NA | NA |
| **Combined comorbidity score, <sup>f</sup> mean (SD)** | 1.0 (2.1) | 1.3 (2.3) | 2.7 (3.3) | 2.1 (2.9) | NA | NA | NA | NA |

Abbreviations: BMI, body mass index (calculated as weight in kilograms divided by height in meters squared); NA, not applicable.
<sup>a</sup> Other/missing sex includes no information, unknown, and other.
<sup>b</sup> Other race includes native Hawaiian or other pacific islander, American Indian or Alaska Native, multiple race, and all other races.
<sup>c</sup> Missing race and ethnicity includes refuse to answer, no information, unknown, and missing values.
<sup>d</sup> For children, obesity was defined as BMI greater or equal to 95th percentile; for adults, obesity was defined as BMI ≥ 30 kg/m<sup>2</sup>.
<sup>e</sup> Smoking status was ascertained based on information in the EHR data. Smoking status was assessed for adults only.
<sup>f</sup>Combined comorbidity score was defined based on Gagne et al.'s combined comorbidity score. Combined comorbidity was assessed for adults only.

Individuals with a positive viral test were older than those with a negative viral test in both age cohorts across most care settings, although non-hospitalized adults who tested positive were younger than those who tested negative (mean age: 49 vs 53 years, P < 0.001). Among both age cohorts, compared to those with a negative viral test, more patients with a positive viral test were Black (26% vs 18% among adults, P < 0.001 and 25% vs 18% among children, P < 0.001) among hospitalized patients and Hispanic (17% vs 10% among adults, P < 0.001 and 23% vs 16% among children, P < 0.001) in both care settings. Adults with a positive viral test were more likely to have obesity in both care settings (5-percent-point difference in both care settings, P < 0.001). Hospitalized children with a positive viral test were more likely to have obesity than those with a negative viral test (21% vs 17%, P = 0.01).

Approximately 35% of hospitalized adults with a positive viral test received dexamethasone, as compared with 21% of hospitalized adults with a negative test (P < 0.001). Among hospitalized patients, individuals with a positive viral test had longer length of stay (mean: 7.3 vs 1.1 among adults, P < 0.001 and 6.2 vs 4.6 among children, P < 0.001) and were more likely to be on mechanical ventilation (5% vs 3% among adults, P < 0.001 and 6% vs 5% among children, P = 0.57) compared to those with negative viral tests in both age cohorts.

### Prevalence of Symptoms Among Children and Adults

Hospitalized patients with a positive viral test had higher prevalence of all symptom outcomes than those with a negative viral test in both adult and youth cohorts 31-150 days after SARS-CoV-2 test (Table 2). Over half (53%) of hospitalized adults with a positive viral test had at least one symptom compared to 44% among those with a negative viral test. Shortness of breath was more prevalent among hospitalized adults who tested positive compared with those who tested negative (17% and 10%, respectively). Similar patterns were observed among children (Table 2). Prevalence of symptoms 31-150 days after SARS-CoV-2 test were similar between non-hospitalized patients testing positive and those testing negative in both age groups (Table 2). The prevalence of symptoms 90-150 days after a SARS-CoV-2 test was lower when compared to symptoms in the 31-150-day period for both children and adults. When comparing patients testing positive to negative, prevalence was similar among non-hospitalized patients but higher among patients testing positive in hospitalized patients (eTable 3). This pattern was consistent among both children and adults.

**Table 2.**
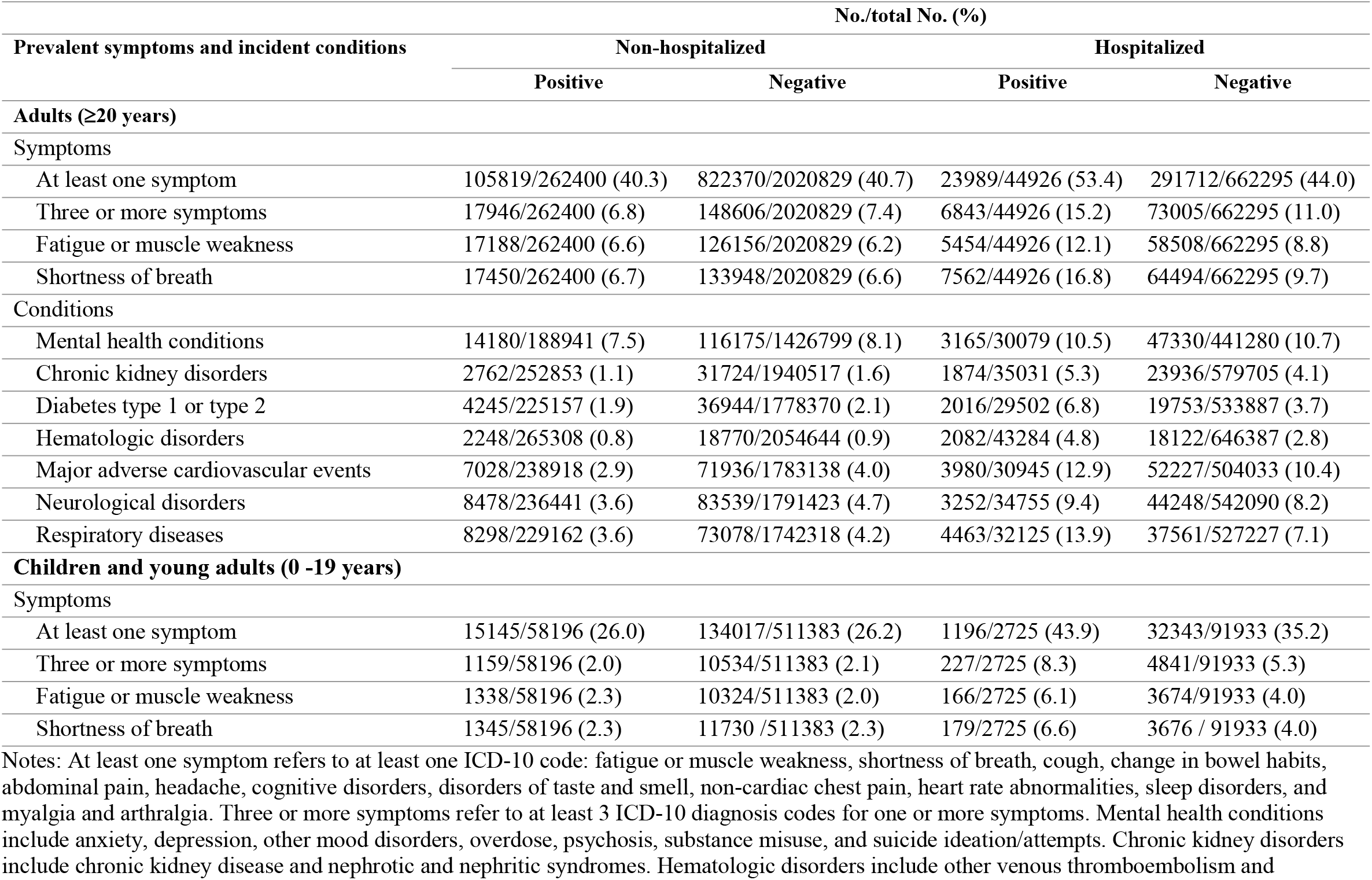
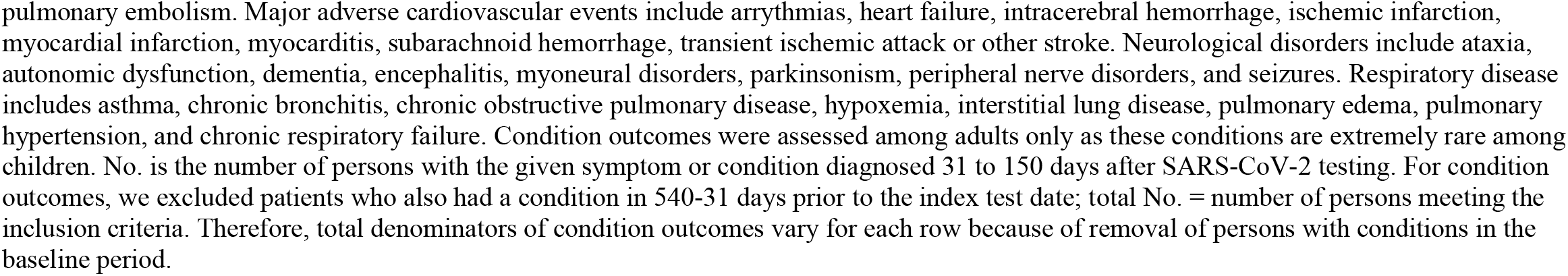
Prevalence of Symptoms and Incidence of Conditions in 31-150 days following SARS-CoV-2 Testing among Adults and Children with Positive and Negative SARS-CoV-2 Test Results.

| Prevalent symptoms and incident conditions | No./total No. (%) |  |  |  |
| --- | --- | --- | --- | --- |
|  | Non-hospitalized |  | Hospitalized |  |
|  | Positive | Negative | Positive | Negative |
| **Adults (≥20 years)** |  |  |  |  |
| Symptoms |  |  |  |  |
| At least one symptom | 105819/262400 (40.3) | 822370/2020829 (40.7) | 23989/44926 (53.4) | 291712/662295 (44.0) |
| Three or more symptoms | 17946/262400 (6.8) | 148606/2020829 (7.4) | 6843/44926 (15.2) | 73005/662295 (11.0) |
| Fatigue or muscle weakness | 17188/262400 (6.6) | 126156/2020829 (6.2) | 5454/44926 (12.1) | 58508/662295 (8.8) |
| Shortness of breath | 17450/262400 (6.7) | 133948/2020829 (6.6) | 7562/44926 (16.8) | 64494/662295 (9.7) |
| Conditions |  |  |  |  |
| Mental health conditions | 14180/188941 (7.5) | 116175/1426799 (8.1) | 3165/30079 (10.5) | 47330/441280 (10.7) |
| Chronic kidney disorders | 2762/252853 (1.1) | 31724/1940517 (1.6) | 1874/35031 (5.3) | 23936/579705 (4.1) |
| Diabetes type 1 or type 2 | 4245/225157 (1.9) | 36944/1778370 (2.1) | 2016/29502 (6.8) | 19753/533887 (3.7) |
| Hematologic disorders | 2248/265308 (0.8) | 18770/2054644 (0.9) | 2082/43284 (4.8) | 18122/646387 (2.8) |
| Major adverse cardiovascular events | 7028/238918 (2.9) | 71936/1783138 (4.0) | 3980/30945 (12.9) | 52227/504033 (10.4) |
| Neurological disorders | 8478/236441 (3.6) | 83539/1791423 (4.7) | 3252/34755 (9.4) | 44248/542090 (8.2) |
| Respiratory diseases | 8298/229162 (3.6) | 73078/1742318 (4.2) | 4463/32125 (13.9) | 37561/527227 (7.1) |
| **Children and young adults (0 -19 years)** |  |  |  |  |
| Symptoms |  |  |  |  |
| At least one symptom | 15145/58196 (26.0) | 134017/511383 (26.2) | 1196/2725 (43.9) | 32343/91933 (35.2) |
| Three or more symptoms | 1159/58196 (2.0) | 10534/511383 (2.1) | 227/2725 (8.3) | 4841/91933 (5.3) |
| Fatigue or muscle weakness | 1338/58196 (2.3) | 10324/511383 (2.0) | 166/2725 (6.1) | 3674/91933 (4.0) |
| Shortness of breath | 1345/58196 (2.3) | 11730 /511383 (2.3) | 179/2725 (6.6) | 3676 / 91933 (4.0) |

Notes: At least one symptom refers to at least one ICD-10 code: fatigue or muscle weakness, shortness of breath, cough, change in bowel habits, abdominal pain, headache, cognitive disorders, disorders of taste and smell, non-cardiac chest pain, heart rate abnormalities, sleep disorders, and myalgia and arthralgia. Three or more symptoms refer to at least 3 ICD-10 diagnosis codes for one or more symptoms. Mental health conditions include anxiety, depression, other mood disorders, overdose, psychosis, substance misuse, and suicide ideation/attempts. Chronic kidney disorders include chronic kidney disease and nephrotic and nephritic syndromes. Hematologic disorders include other venous thromboembolism and
pulmonary embolism. Major adverse cardiovascular events include arrhythmias, heart failure, intracerebral hemorrhage, ischemic infarction, myocardial infarction, myocarditis, subarachnoid hemorrhage, transient ischemic attack or other stroke. Neurological disorders include ataxia, autonomic dysfunction, dementia, encephalitis, myoneural disorders, parkinsonism, peripheral nerve disorders, and seizures. Respiratory disease includes asthma, chronic bronchitis, chronic obstructive pulmonary disease, hypoxemia, interstitial lung disease, pulmonary edema, pulmonary hypertension, and chronic respiratory failure. Condition outcomes were assessed among adults only as these conditions are extremely rare among children. No. is the number of persons with the given symptom or condition diagnosed 31 to 150 days after SARS-CoV-2 testing. For condition outcomes, we excluded patients who also had a condition in 540-31 days prior to the index test date; total No. = number of persons meeting the inclusion criteria. Therefore, total denominators of condition outcomes vary for each row because of removal of persons with conditions in the baseline period.

### Association between SARS-CoV-2 Infection and Prevalent Symptoms 31 to 150 Days After Testing among Hospitalized Children and Adults

Hospitalized adults with a positive viral test had increased odds of being diagnosed with at least one symptom (adjusted odds ratio [aOR], 1.17[95% CI, 1.11-1.23]), three or more symptoms (aOR, 1.16[95% CI, 1.08 – 1.26]), fatigue (aOR, 1.12[95% CI, 1.05 – 1.18]), and shortness of breath (aOR, 1.50[95% CI, 1.38-1.63]) 31 to 150 days after SARS-CoV-2 test (Figure 1). We found similar associations between SARS-CoV-2 infection and symptoms 90-150 days after testing among hospitalized adults (≥1 symptom: aOR, 1.15[95% CI, 1.09-1.22]); three or more symptoms: aOR, 1.23[95% CI, 1.14-1.32]. eFigure 1).

**Figure 1.**
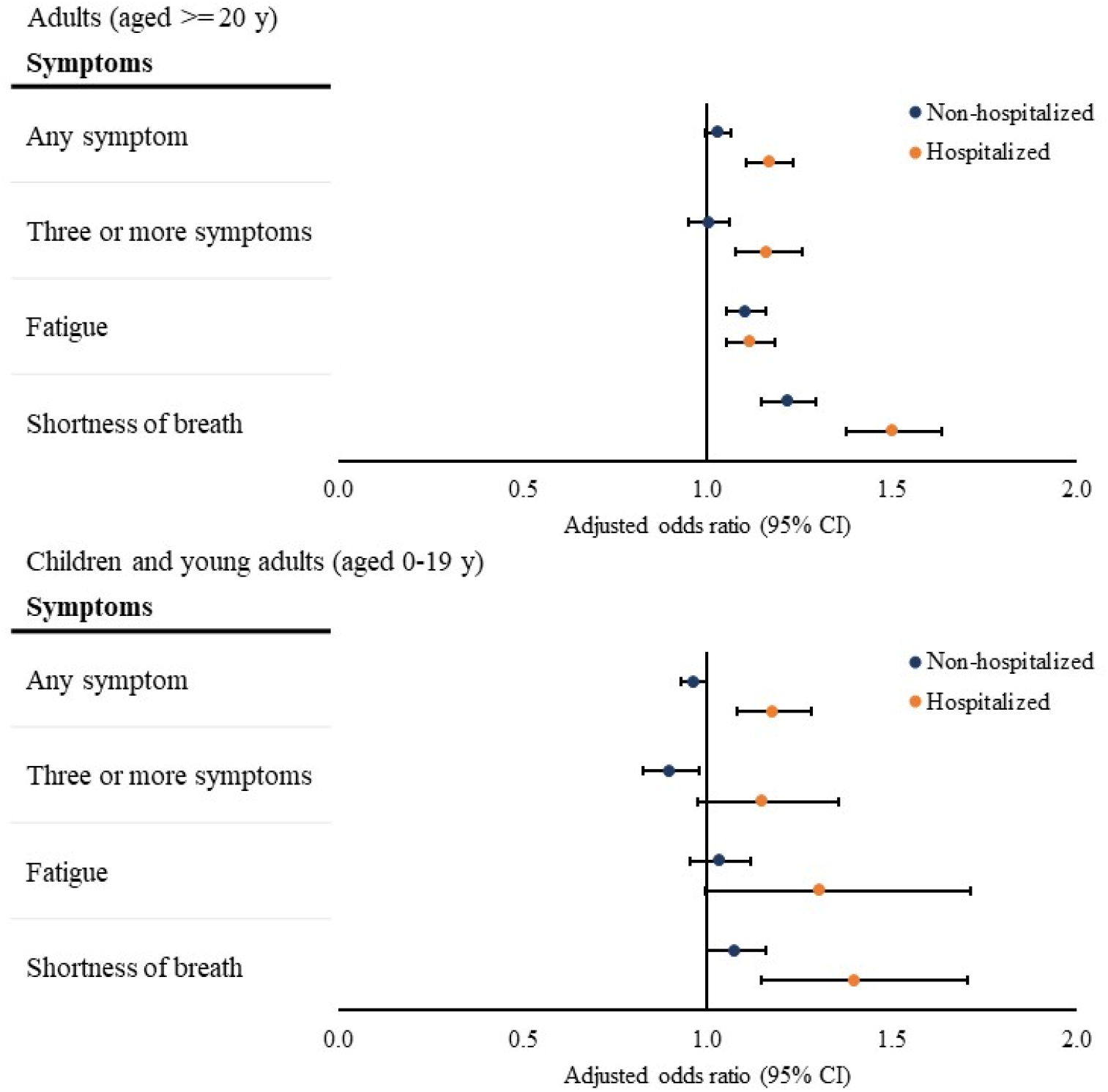
Association between SARS-CoV-2 Infection and Symptoms in 31 to 150 Days After SARS-CoV-2 Testing. Notes: Associations were assessed by comparing the presence of each outcome between patients with a positive viral test and those with a negative viral test, adjusting for baseline demographic and clinical characteristics as confounders using logistic regressions. The overall odds ratios were calculated using meta-analyses from site-specific estimates. At least one symptom refers to at least one symptom among fatigue or muscle weakness, shortness of breath, cough, change in bowel habits, abdominal pain, headache, cognitive disorders, disorders of taste and smell, non-cardiac chest pain, heart rate abnormalities, sleep disorders, and myalgia and arthralgia. Three or more symptoms refer to at least 3 different ICD-10 diagnosis codes for one or more symptoms.

Hospitalized children with a positive viral test had increased odds of being diagnosed with at least one symptom (aOR, 1.18[95% CI, 1.08-1.28]) and shortness of breath (aOR, 1.40[95% CI, 1.15-1.70]) 31-150 days after SARS-CoV-2 test (Figure 1). The odds of being diagnosed with three or more symptoms (aOR, 1.15[95% CI, 0.98 -1.36]) or fatigue (aOR, 1.31[95% CI, 0.99 -1.71]) did not differ between hospitalized children with a positive and a negative viral test (Figure 1). We found similar associations between SARS-CoV-2 infection and symptoms 90-150 days after SARS-CoV-2 test among hospitalized children (≥1 symptom: aOR, 1.16[95% CI, 1.03-1.31]; three or more symptoms: aOR, 1.22[95% CI, 0.95-1.55]. eFigure 1).

### Association between SARS-CoV-2 Infection and Prevalent Symptoms 31 to 150 Days After Testing among Non-hospitalized Children and Adults

Among non-hospitalized adults, those with a positive viral test had higher odds of being diagnosed with fatigue (aOR, 1.11[95% CI, 1.05-1.16]) and shortness of breath (aOR, 1.22[95% CI, 1.15-1.29]) 31-150 days after the index date compared with those with a negative viral test (Figure 1). The odds of being diagnosed with at least one symptom (aOR, 1.03[95% CI, 1.00 -1.06]) or three or more symptoms (aOR, 1.00[95% CI, 0.95 -1.06]) were similar between non-hospitalized adults with a positive and a negative viral test (Figure 1). Results were similar for symptoms 90-150 days after testing (≥1 symptom: aOR, 1.00[95% CI, 0.97-1.04]; three or more symptoms: aOR, 1.01[95% CI, 0.96-1.07]. eFigure 1).

Among non-hospitalized children, those with a positive viral test had a decreased odds of being diagnosed with three or more symptoms (aOR, 0.90[95% CI. 0.83-0.98]) 31 to 150 days after the index date when compared with those with a negative viral test. The odds of being diagnosed with at least one symptom, fatigue, and shortness of breath were all similar between those with a positive and those with a negative viral test among non-hospitalized children (Figure 1). Similar results were found for symptoms 90-150 days after testing (≥1 symptom: aOR, 0.95[95% CI, 0.92-0.98]; three or more symptoms: aOR, 0.85[95% CI, 0.77-0.94]. eFigure 1).

### Incidence of New Conditions Among Adults

Hospitalized adults with a positive viral test had higher incidence of almost all conditions, except for mental health conditions, compared with those with negative viral test (Table 2). Hospitalized adults with a positive viral test were most likely to be newly diagnosed with respiratory diagnoses (14%) when compared with those with a test-negative patients (7%). The incidence of respiratory diseases was primarily driven by chronic respiratory failure (8%) and hypoxemia (4%) among hospitalized patients with a positive viral test. Non-hospitalized adults with a positive viral test had approximately similar incidence of type 1 or type 2 diabetes (2%), hematologic disorders (1%), mental health conditions (8%), and respiratory diseases (4%), and had lower incidence of chronic kidney disorders (1% vs 2%), major adverse cardiovascular events (3% vs 4%), and neurological disorders (4% vs 5%), than those who had a negative viral test (Table 2).

### Association between SARS-CoV-2 Infection and New Conditions among Adults in 31 to 150 Days After Testing

The risk of being newly diagnosed with type 1 or type 2 diabetes (adjusted hazard ratio [aHR], 1.25[95% CI, 1.17-1.33]), hematologic disorders (aHR, 1.19[95% CI, 1.11-1.28]), and respiratory disease (aHR, 1.44[95% CI, 1.30-1.60]) were higher among hospitalized adults with a positive viral test compared with those with a negative viral test (Figure 2), whereas the risk of being newly diagnosed with mental health conditions (aHR, 0.85[95% CI, 0.80-0.90]), major adverse cardiovascular events (aHR, 0.91[95% CI, 0.83-0.99]), and neurological disorders (aHR, 0.89[95% CI, 0.85-0.94]) was lower among hospitalized adults with a positive viral test relative to those with a negative viral test (Figure 2). The risk of being newly diagnosed with chronic kidney disorders was not statistically different between hospitalized adults with a positive and a negative viral test (Figure 2).

**Figure 2.**
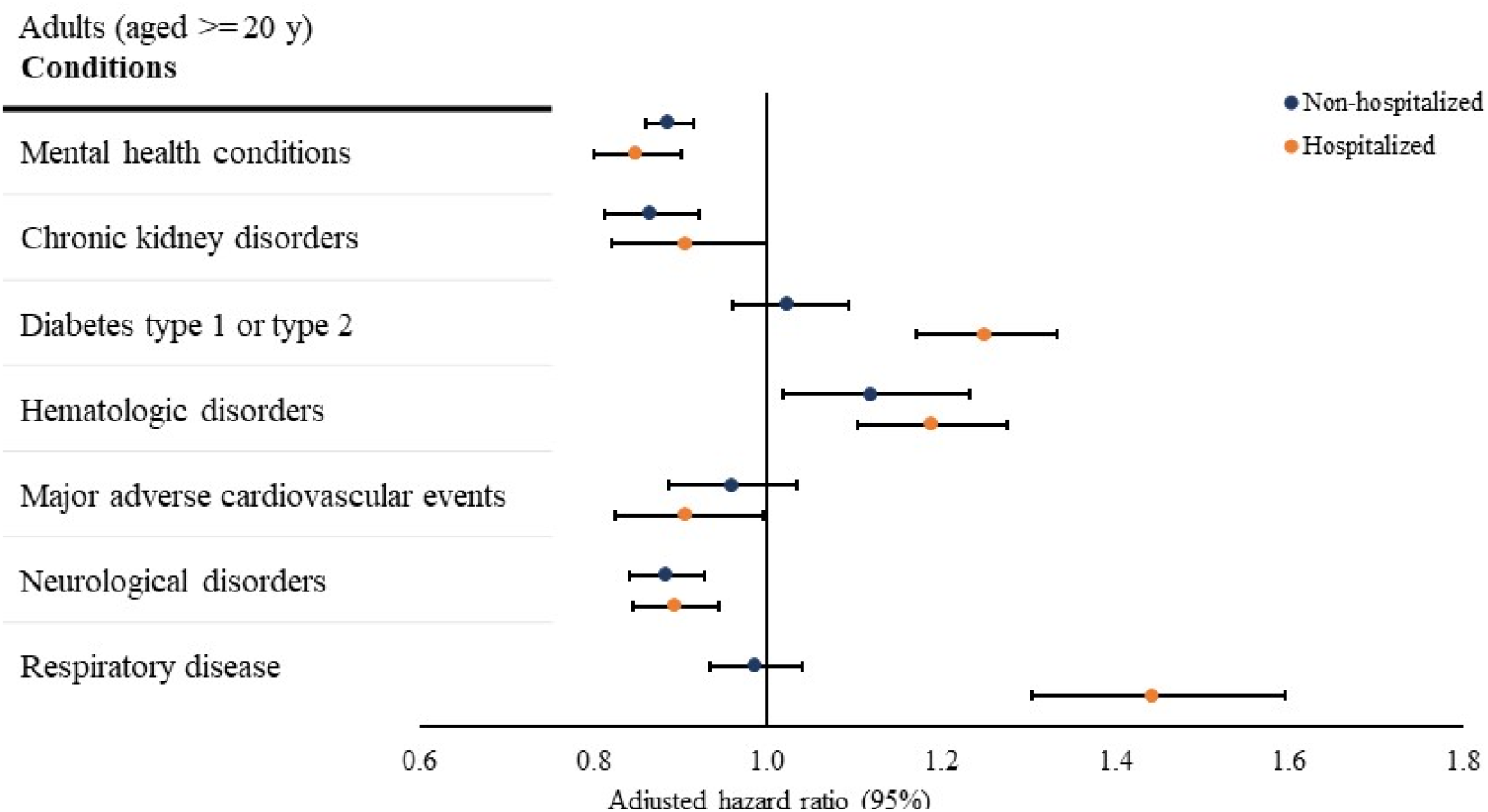
Association between SARS-CoV-2 Infection and Conditions in 31 to 150 Days After SARS-CoV-2 Testing. Notes: Associations were assessed using Cox proportional hazard regression models, accounting for time from the beginning of the post-acute period (31 days post) to the earliest presence of the first diagnostic code for each condition (event) and the end of the outcome period (150 days post-censoring). The overall hazard ratios were calculated using meta-analyses from site-specific estimates. Mental health conditions include anxiety, depression, other mood disorders, overdose, psychosis, substance misuse, and suicide ideation/attempts. Chronic kidney disorders include chronic kidney disease and nephrotic and nephritic syndromes. Hematologic disorders include other venous thromboembolism and pulmonary embolism. Major adverse cardiovascular events include arrythmias heart failure, intracerebral hemorrhage, ischemic infarction, myocardial infarction, myocarditis, subarachnoid hemorrhage, transient ischemic attack or other stroke. Neurological disorders include ataxia, autonomic dysfunction, dementia, encephalitis, myoneural disorders, parkinsonism, peripheral nerve disorders, and seizures. Respiratory disease includes asthma, chronic bronchitis, chronic obstructive pulmonary disease, hypoxemia, interstitial lung disease, pulmonary edema, pulmonary hypertension, and chronic respiratory failure.

Among non-hospitalized adults, those with a positive viral test had an increased risk of being newly diagnosed with hematologic disorders (aHR, 1.12[95% CI, 1.02-1.23]) and a decreased risk of being newly diagnosed with mental health conditions (aHR, 0.89[95% CI, 0.86-0.92]), chronic kidney disorders (aHR, 0.87[95% CI, 0.81-0.92]), and neurological disorders (aHR, 0.89[95% CI, 0.84-0.93]), compared with non-hospitalized adults with a negative viral test (Figure 2). The risks of being newly diagnosed with type 1 or type 2 diabetes, major adverse cardiovascular events, and respiratory disease were not statistically different between non-hospitalized adults with a positive and a negative viral test (Figure 2).

## Discussion

Using longitudinal EHR data of 3.7 million individuals who were tested for SARS-CoV-2 infection and received care from health systems associated with 43 PCORnet sites across the U.S., we identified that adults with a positive SARS-CoV-2 test were at increased odds of being diagnosed with certain symptoms (e.g., fatigue and shortness of breath) and were at a higher risk of being newly diagnosed with certain conditions (e.g., diabetes and hematologic disorders) as potential PASC 31-150 days after testing, compared with patients who always tested negative for SARS-CoV-2. Hospitalized children with a positive SARS-CoV-2 test also were at increased odds of being diagnosed with symptoms, including shortness of breath, compared to those hospitalized children testing negative. The comprehensive and longitudinal information from EHRs enabled us to adjust for various symptoms and conditions that patients had before SARS-CoV-2 infection.

We found that differences in prevalence of symptoms and incidence of conditions following SARS-CoV-2 positive and negative test results were more evident among hospitalized patients than non-hospitalized patients. After adjusting for confounders, hospitalized adults testing positive had increased odds of having all symptom outcomes and increased risk of being newly diagnosed with type 1 or 2 diabetes, hematologic disorders, and respiratory diseases compared with hospitalized adults testing negative. Hospitalized children who were positive had increased odds of having at least one symptom and shortness of breath, relative to hospitalized children who were negative. These findings are consistent with literature reports showing that patients with more severe acute SARS-CoV-2 infection (i.e., hospitalized patients) have a higher risk of developing PASC conditions and symptoms [36, 37].

We found relatively small differences in symptoms and conditions between non-hospitalized patients who tested positive and those who tested negative. Regression analyses adjusting for confounders suggested that non-hospitalized adults testing positive had higher odds of being diagnosed with fatigue and shortness of breath and higher risk of being newly diagnosed with hematologic disorders compared with those testing negative. We found no symptoms with higher odds among non-hospitalized children testing positive compared with those testing negative. This evidence adds to the growing but still limited literature on post-acute sequelae of SARS-CoV-2 infection among non-hospitalized patients.

We did find some conditions that were more common among hospitalized adults testing negative; these included mental health conditions, major cardiovascular events, and neurologic disorders. While it is possible that these conditions are less common after SARS-CoV-2 infection, these differences also might reflect conditions for which patients testing negative were hospitalized. We could not define the primary reason for hospitalizations and thus could not control for the possibility that patients may have been hospitalized for conditions that persisted in the post-acute period of 31 to 150 days after index date. We did restrict these analyses to those patients who did not have these conditions during the baseline period.

Several prior studies have examined incidence of conditions and symptoms after SARS-CoV-2 infection compared with control groups that did not have COVID-19 [3]. In a study of patients 65 years or older participating in US Medicare Advantage plans, researchers compared the incidence of 18 conditions after a positive test for SARS-CoV-2 or a diagnosis of COVID-19 (N=87,337), compared to several populations that did not have documented COVID-19: a 2020 group (87,337), a 2019 group (88,070), and a group with documented viral lower respiratory tract diagnoses before 2020 (73,490). Respiratory failure, fatigue, memory difficulties, kidney injury, hypercoagulability and cardiac rhythm disorders had the highest incidences among patients with COVID-19 relative to comparator populations, with higher relative differences in incidences for those who had severe COVID-19 disease. Our study examined both symptoms and conditions and found a smaller list of conditions associated with SARS-CoV-2 infection compared to contemporaneous controls of those testing negative.

Another study among US veterans compared non-hospitalized patients with a positive viral test (N= 73,435) with non-hospitalized patients without a positive viral test (4,990,835) [1]. In the study, COVID-19 patients had increased risk of developing incident conditions including respiratory conditions, diseases of the nervous system, mental health conditions, cardiovascular conditions, among other conditions, and incident symptoms like malaise and fatigue in the first 30 days after a positive test. A comparison between hospitalized COVID-19 patients (13,654) with patients who were hospitalized for influenza (13,997, identified during October 1, 2016– February 29, 2020) found that hospitalized COVID-19 patients had increased risks of being diagnosed with incident conditions including neurological disorders, mental health disorders, cardiovascular disorders, among other conditions, 30 days after hospital admission. Patients in this study were predominantly male (88%), relatively older (mean age 59 years), and White (70%). Our study focused on a more generalizable population that included both adults and children and compared hospitalized COVID-19 patients with those hospitalized for all other conditions.

Another population-based study examined 27 conditions and symptoms recorded in hospital inpatient and outpatient settings from 2 weeks to 6 months after a SARS-CoV-2 test among non-hospitalized COVID-19 patients (N = 8,983) compared with non-hospitalized patients without COVID-19 (80,894) in Denmark [38]. The study found that non-hospitalized COVID-19 patients had increased risks for dyspnea and venous thromboembolism following SARS-CoV-2 infection. This study included patients in the early waves of the pandemic (February 27 to May 31, 2020) and was among a few studies including both adults and children (7% under 18 years). However, their analysis did not stratify by age.

Our study provides evidence for symptoms and conditions as potential long-term sequelae of SARS-CoV-2 infection for children and adults, especially among those with a hospitalization associated with SARS-CoV-2 test. These results have important clinical and public health implications. Clinicians and public health agencies should monitor for the development and persistence of symptoms and conditions after COVID-19, especially among those who are hospitalized. The higher burden of PASC symptoms and conditions post-COVID also should encourage investment in clinical and public health resources needed to deliver care to treat and prevent PASC, including ongoing support for trials underway to evaluate effectiveness of treatments for specific post-COVID conditions [39]. Trials might be more impactful if they focused on patients initially hospitalized for COVID-19, because of the higher incidence among these patients.

This study is subject to several limitations. First, use of EHR data to ascertain symptoms and conditions may have led to an underestimation of real prevalence and incidence as we only observe diagnosis codes for these symptoms and conditions when patients have a clinical encounter with health systems of participating sites. Symptoms may be much less likely to be entered into the EHR as diagnostic codes; clinicians often describe symptoms only in unstructured notes. This is particularly an issue among patients with limited healthcare access. Similarly, patients who always tested negative might have had a positive test that was not captured in EHR (e.g., self-test at home). Thus, it is possible that some patients in the control group may have tested positive at some point, perhaps within the follow-up period of 30-150 days after their negative test. This differential misclassification would bias results toward the null. Second, we defined symptoms or conditions as the occurrence of one ICD-10-CM diagnostic code 31 to 150 days following SARS-CoV-2 infection. This approach was used to enhance sensitivity for detection of possible SARS-CoV-2 sequelae in the short interval of 31 to 150 days after the test but may have lower specificity. Although using 2 or more occurrences of diagnostic codes may have enhanced specificity, this would have restricted the study to patients with multiple encounters in the same site, such as sicker patients or patients with better access to healthcare. Third, although we have adjusted for a comprehensive set of confounders in regression models, certain important covariates, such as vaccination status, were not included due to data limitations. Vaccination data is often missing in EHR systems for most health systems because of incomplete capture of data from state immunization registries. Fourth, we used hospitalization within 16 days of a positive test for SARS-CoV-2 infection as a proxy for COVID-19 severity, which may have resulted in misclassification if patients with a positive test were hospitalized for reasons other than acute COVID-19 illness. Fifth, we were unable to ascertain whether SARS-CoV-2 testing was conducted because of symptoms, as a part of routine surveillance, or for travel purposes. Hospitalized persons who tested negative for SARS-CoV-2 included those hospitalized for nonviral illness (e.g., pregnancy, trauma, chronic conditions, elective procedures) and may have biased our estimates if these illnesses were associated with conditions or symptoms assessed in this study. There may be multiple etiologies for symptoms and conditions examined in this report; the same is true for patients tested in the non-hospital setting prior to certain tests or procedures that were for illnesses that also might have been associated with the conditions or symptoms assessed in this study. Future studies may compare patients hospitalized for SARS-CoV-2 infection only with patients hospitalized for influenza and other lower respiratory tract illnesses. Finally, for covariates with missing values (e.g., sex and race), we adjusted for missing values as a separate category in the analyses. Imputing missing values may be a more robust approach.

In conclusion, we examined associations between SARS-CoV-2 infection and a set of symptoms and conditions as potential post-acute sequelae of SARS-CoV-2 infection among both hospitalized and non-hospitalized adults and children. Our findings suggest an association of post-acute sequelae of SARS-CoV-2 infection with higher severity of acute SARS-CoV-2 infection and highlight certain symptoms and conditions that are more common among patients testing positive for SARS-CoV-2. Future research is warranted to examine prevention and treatment of these symptoms and conditions to help patients recover from SARS-CoV-2 infection.

## Data Availability

Data cannot be shared publicly because of the use of the PCORnet data is under specific data use agreement, which requires IRB approval and PCROnet's review and approval. Data are available from the National Patient-Centered Clinical Research Network (https://pcornet.org/) for researchers who meet the criteria for access to confidential data. Researchers who are interested in using PCORnet data can request data via the established process. https://pcornet.org/front-door/

## Code Availability

All codes used for this query are available at GitHub at https://github.com/PCORnet-DRN-OC/Query-Details/blob/master/Long%20COVID%20Symptoms/CodeList_Long_COVID_Symptoms_Analytic_Query2_v1.0.xlsx

## Collaborative Authors: PCORnet® Network Partners

Faraz S. Ahmad, MD, MS, Northwestern University Feinberg School of Medicine; H. Timothy Bunnell, PhD, Nemours Children’s Health; Olveen Carrasquillo, MD, MPH, University of Miami Miller School of Medicine; Elizabeth A. Chrischilles, PhD, The University of Iowa; Dimitri A. Christakis, MD, MPH, Seattle Children’s Research Institute; Bernard P. Chang, MD, PhD, Columbia University Irvine Medical Center; Janis L. Curtis, MSPH, MA, Duke University School of Medicine; Soledad A. Fernandez, PhD, The Ohio State University; Christopher B. Forrest, MD, PhD, Children’s Hospital of Philadelphia; Daniel Fort, PhD, MPH, Ochsner Clinic Foundation; David A Hanauer, MD, MS, University of Michigan; Rachel Hess, MD, MS, University of Utah; Benjamin D. Horne, PhD, MStat, MPH, Intermountain Healthcare; Philip Giordano, MD, FACEP, Orlando Health, Inc.; William Hogan, MD, MS, University of Florida; Kenneth H. Mayer, MD, Fenway Health; Abu Saleh Mohammad Mosa, PhD, MS, FAMIA, University of Missouri; James C. McClay, MD, University of Nebraska Medical Center; Samyuktha Nandhakumar, MS, UNC School of Medicine; Bridget Nolan, Department of Population Medicine, Harvard Pilgrim Health Care Institute; Jihad S. Obeid, MD, FAMIA, Medical University of South Carolina; Brian Ostasiewski, Wake Forest School of Medicine; Anuradha Paranjape, MD, MPH, Lewis Katz School of Medicine at Temple University; Lav Patel, BS, MS, University of Kansas Medical Center; Suchritra Rao, MD, Children’s Hospital Colorado; Patricia S. Robinson, PhD, APRN, AdventHealth Research Institute; William E. Trick, MD, Cook County Health; Jonathan C. Silverstein, MD, MS, University of Pittsburgh School of Medicine

